# Comparison of causes of stillbirth and child deaths as determined by verbal autopsy and minimally invasive tissue sampling

**DOI:** 10.1101/2024.03.11.24304131

**Authors:** Nega Assefa, Anthony Scott, Lola Madrid, Merga Dheresa, Gezahegn Mengesha, Shabir Mahdi, Sana Mahtab, Ziyaad Dangor, Nellie Myburgh, Lesego Kamogelo Mothibi, Samba O. Sow, Karen L. Kotloff, Milagritos D. Tapia, Uma U. Onwuchekwa, Mahamane Djiteye, Rosauro Varo, Inacio Mandomando, Ariel Nhacolo, Charfudin Sacoor, Elisio Xerinda, Ikechukwu Ogbuanu, Solomon Samura, Babatunde Duduyemi, Alim Swaray-Deen, Abdulai Bah, Shams El Arifeen, Emily S Gurley, Mohammed Zahid Hossain, Afruna Rahman, Atique Iqbal Chowdhury, Bassat Quique, Portia Mutevedzi, Argeseanu Solveig, Dianna Blau, Cyndy Whitney

**Affiliations:** College of Health and Medical Sciences, Haramaya University, Harar, Ethiopia; London School of Hygiene and Tropical Medicine, London, United Kingdom; South African Medical Research Council Vaccines and Infectious Diseases Analytics Research Unit, University of the Witwatersrand, Johannesburg, South Africa; Centre pour le Développement des Vaccins (CVD-Mali), Ministère de la Santé, Bamako, Mali; Department of Pediatrics, Center for Vaccine Development and Global Health, University of Maryland School of Medicine, Baltimore, Maryland, USA; ISGlobal, Hospital Clínic, Universitat de Barcelona, Barcelona, Spain; Centro de Investigação em Saúde de Manhiça (CISM), Maputo, Mozambique; Instituto Nacional de Saude, Ministerio de Saude, Maputo, Mozambique; Crown Agents, Freetown, Sierra Leone; World hope international, Makeni, Sierra Leone; University of Sierra Leone Teaching Hospitals Complex, Sierra Leone; University of Ghana Medical School; FOCUS 10000, Freetown, Sierra Leone; Maternal and Child Health Division, icddr,b, Dhaka, Bangladesh; Bloomberg School of Public Health, Johns Hopkins University, Baltimore, Maryland, United States of America; International Centre for Diarrhoeal Disease Research Bangladesh (icddr,b), Dhaka, Bangladesh; PEI, Infectious Disease Division, icddr,b, Dhaka, Bangladesh; Global Health Institute, Emory University, Atlanta, Georgia, United States of America; Global Health Center, Centers for Disease Control and Prevention, Atlanta, Georgia, United States of America

**Author notes:** **Corresponding author:** Nega Assefa, College of Health and Medical Sciences, Haramaya University, Harar Campus, SGS building, Ethiopia. These authors contributed equally to this work (Dianna Blau, Cyndy Whitney).

**Keywords:** Cause of death, Determination of Cause of Death, CHAMPS, sub-Saharan Africa, Open VA, InterVA-4 package

## Abstract

**Background:** In resource-limited settings where vital registration and medical death certificates are unavailable or incomplete, verbal autopsy (VA) is often used to attribute causes of death (CoD), identify the distribution and trends of diseases, and prioritize resource allocation and interventions. However, VA findings can be non-specific, as this tool is based on family members’ recall of symptoms rather than objective diagnostic testing. We aimed to compare the CoD diagnoses obtained in stillbirths and children below five years of age (<5s) through two very different approaches; namely: 1) VA; and 2) the results obtained through the use of Minimally Invasive Tissue Sampling (MITS) and rigorous diagnostic testing, as part of the approach proposed by the Child Health and Mortality Prevention Surveillance (CHAMPS).

**Methods:** CHAMPS identified stillbirths and deceased children <5s in real time between 2017 and 2021 in catchment areas in seven low- and middle-income countries (LMICs): Bangladesh, Ethiopia, Kenya, Mali, Mozambique, Sierra Leone, and South Africa. Deaths were eligible for MITS if identified <24 hours after death, legal concerns were not present, burial had not occurred, and parents consented. CHAMPS teams utilized information from MITS and VA to determine the causes of death (CoDs); if not eligible for MITS, the InterVA software utilized only VA information to determine the CoDs. CHAMPS attributed CoD using expert panels that reviewed clinical evidence microbiological, and histopathological results from MITS to derive the CoDs (Determination of Cause of Death [DeCoDe]). The InterVA4 package of OpenVA software automatically assigned the underlying CoDs using the Bayesian probabilistic modeling technique. These automatically assigned CoDs from OpenVA were compared to the gold-standard of the CHAMPS-attributed CoDs to evaluate both systems’ agreement, weaknesses, and strengths using Lin’s concordance correlation coefficient.

**Results:** Data from 2852 deaths that underwent MITS were analysed. The most common age categories were stillbirths (n=1075, 37.7%) and neonatal deaths (n=1077, 37.8%). Overall concordance of InterVA4 and DeCoDe in assigning causes of death across surveillance sites, age groups, and causes of death was poor (0.75 with 95% CI: 0.73 – 0.76) and lacked precision. We found substantial differences in agreement among surveillance sites, with Mali showing the lowest and Mozambique and Ethiopia the highest concordance. Lin’s concordance correlation coefficient for children aged < 1 year was 0.69 (95%CI: 0.65 – 0.71), and for children aged 1-4 years was 0.28 (95%CI: 0.19 – 0.37)

**Conclusion:** The InterVA4 assigned CoD agrees poorly in assigning causes of death for under-fives and stillbirths. Because VA methods are relatively easy to implement, such systems could be more useful if algorithms were improved to more accurately reflect causes of death, for example, by calibrating algorithms to information from programs that used detailed diagnostic testing to improve the accuracy of COD determination.

## Introduction

In contrast to countries in high-income countries, most low and middle-income countries (LMIC) that have higher rates of mortality among children lack adequate systematic mortality surveillance [1]. For example, death registration coverage varies from nearly 100% in the WHO European region to less than 10% in the WHO African region [2]. In LMICs, deaths are often not attended by health professionals, not medically certified, not recorded in a timely way, and, even when recorded, the information is stored inappropriately [3]. LMICs also do not have the infrastructure or resources to establish and maintain data systems that conclusively identify causes of death in their populations [4]. Not having appropriate legislation or health policies on data systems compounds these challenges, leading to ineffective formulation and implementation of interventions to reduce mortality at a population level [5, 6].

One relatively simple method to try to identify the CoDs is through a verbal autopsy (VA) [7–10]. To conduct a VA, workers who are trained on the method but who typically lack specific clinical training interview family members or caregivers of the deceased using a structured questionnaire; the workers also solicit a complete narrative of the circumstances surrounding death [11]. Causes of death (CoDs) can then be easily generated from the structured questionnaire responses by probabilistic analytic algorithms that are freely available and accessible online. Because other modalities, such as physician coding, may not be efficient, affordable, and sustainable in resource-limited settings, publicly available CoDs generating software from the VA has become an essential public health tool for mortality estimation and identifying population-level CoDs in resource-limited settings [12, 13].

However, VA has several weaknesses [14, 15]. The method can produce conflicting and unreliable CoD results because it relies on the quality and accuracy of information provided by family members who typically lack clinical training. The community’s sociocultural and recall biases can affect families’ responses. The VA forms do not collect information on known or pre-existing medical conditions determined based on diagnostic testing, as the families are the proxy respondents who might not have access to past clinical information [16–18]. The presence of multiple VA algorithms and the tool’s inherent limitations with accurately determining pre-existing or new medical problems make it challenging to understand and determine the CoD for conditions with complex cause-of-death pathways or highly non-specific signs and symptoms [19, 20]. In addition, the VA does not generate the complete mortality pathways, such as the immediate or morbid pathways, but only determines the underlying CoDs with various probabilistic scores, which may not adequately capture complicated medical histories. For stillbirths, the VA only describes the body’s condition, which has been shown to not accurately reflect or even impossible to determine the causes of death among stillbirths [21, 22]

The Child Health and Mortality Prevention Surveillance (CHAMPS) is a collaborative network in sub-Saharan Africa and South Asia that uses other additional approaches, including Minimally Invasive Tissue Sampling (MITS), clinical data, histopathological and microbiological findings, and the VA narrative itself, to provide reliable, detailed, and specific causes of stillbirths and child deaths [23–25]. With the support of the Bill & Melinda Gates Foundation, CHAMPS was launched in 2017 in several high child-mortality countries to provide reliable data on cause-specific mortality, which is fundamental to evidence-based health policy and public health interventions [26]. CHAMPS uses thorough postmortem diagnostic testing along with some parts of the VA (open narrative and raw answers to VA questionnaire, but not the VA-derived diagnoses) and review of clinical records. A local panel of experts reviews all the information package and assigns underlying, intermediate, and immediate causes of death, a process called Determination of Cause of Death (DeCoDe). While CHAMPS’ methods produce high-quality cause-of-death information for those children evaluated, the postmortem diagnostic testing protocols require rapid death identification and collection of specimens before a child is buried. Therefore, many deaths in CHAMPS catchment areas do not undergo such testing; in contrast, evaluation of deaths using VA can be done after burial, at a family’s convenience, which is usually 2 to 12 weeks [27].

This study compares the type, quality, and amount of CoD information generated from “VA only” and CHAMPS’s methods; we also assess the concordance of the results generated from the two approaches. While CHAMPS generates specific microbiology and pathology diagnoses, this study focuses on the accuracy of the CoDs assigned by both systems. Methodologically speaking, although VA data was taken using the WHO 2016 questionnaire, the ICD-10 diagnosis determined by the CHAMPS methods was mapped to syndromic categories from the 2016 WHO Verbal Autopsy guideline for comparative purposes. By analysing data from CHAMPS sites in seven countries in Sub-Saharan Africa and Southeast Asia (Bangladesh, Ethiopia, Kenya, Mali, Mozambique, Sierra Leone, and South Africa), we aimed to inform public health leaders and policymakers of the strengths and weaknesses, relevance, and consistency of these approaches.

## Methods

### Study settings and design

The CHAMPS Network longitudinally collects robust and standardized data from its sites to understand and track preventable causes of childhood death in high-mortality areas. The CHAMPS network details have been published elsewhere. [28–30]. All CHAMPS network sites are in research centers with pre-existing Demographic Health Surveillance Systems (HDSS) or have built capacity to closely follow up their catchment’s population, enabling them to capture periodic mortality surveillance data. An HDSS is an open, dynamic cohort that follows residents of a geographically defined area over time, tracking the occurrence of births, deaths, marriages, pregnancies, and migrations. HDSS teams in CHAMPS sites enumerate these events during routine household visits. To identify stillbirths and deaths in children under five as soon as they occur, mortality surveillance in CHAMPS sites also involves community informants, healthcare workers, and other methods.

### Data collection

CHAMPS study procedures have been published elsewhere [31]. Briefly, data from deaths identified through HDSS and mortality surveillance are collected longitudinally from notified deaths in the communities and health facilities within the catchment. An <5s or stillbirth identified within 24 hours of death or 72 hours if refrigerated whose family had been living in the respective catchment area for at least four to six months is eligible to be enrolled for the CHAMPS and requested to provide consent for Minimally Invasive Tissue Sampling (MITS). MITS includes postmortem collection of swabs, postmortem biopsies of vital organs, and body fluids for cytopathologic and microbiologic examination. Clinical information found at the health facilities and the community where the stillbirths and <5s deaths occurred is also collected, and families of the deceased are interviewed using the VA questionnaire.

### Verbal autopsy questionnaire

All sites use the WHO-2016 VA questionnaire, customized to include content enhancements, skip logic, and unit of measurement corrections for the CHAMPS study [32]. VA questionnaires were translated into local languages and collected information on age, sex, place of death, and symptoms observed during the late-life period of the deceased. The questionnaire also contains the symptom duration checklist, which is arranged loosely around anatomical systems and is intended to be informative in leading to a diagnosis of probable CoDs and narrowing the number of possible differential diagnoses.

### Causes of Death assignment from VA

We used the InterVA-4 package from Open-VA to auto-generate the cause for each enrolled death at the surveillance site [33, 34]. Open-VA uses Bayesian probabilistic modeling to assign likelihoods to causes of death based on coded responses to verbal autopsy questionnaires and ascribes corresponding ICD-10 codes [35, 36]; InterVA-4 algorithms do not consider information in the narrative section of the VA. This system mainly generates one likely CoDs and, if a single cause is not clear, three causes with probability values. The generated CoD with the highest probability was considered the underlying cause for comparison with the CoD assigned by CHAMPS DeCoDe.

### Determination of Cause-of-Death (DeCoDe) using Minimally Invasive Tissue Sampling (MITS)

Following the World Health Organization (WHO) application of the International Classification of Diseases – Version 10 (ICD-10), the DeCoDe expert panel determined the underlying cause and, for some deaths, one or more intermediate causes and an immediate CoD [35, 37]. Because we also compared the immediate CoDs assigned by the DeCoDe with the InterVA4’s underlying CoDs, as the InterVA4 does not designate immediate CoDs as the DeCoDe and found no significant difference, we decided Only to use the underlying CoDs assigned by DeCoDe for comparison purposes.

To ensure the DeCoDe panel is aligned with the correct diagnoses and decrease bias, all panel members of each surveillance site follow a standard operating procedure and CHAMPS Diagnosis Standards [38].

The assigned causes of death by the DeCoDe panel were converted and categorized to the corresponding VA diagnosis using the 2016 WHO VA category definitions of the verbal autopsy standard [39]. The standard has a conversion table that shows and defines the VA diagnosis category and title with its corresponding ICD-10 codes. This conversion and categorization enable comparison of the generated CoD InterVA4 with the DeCoDe, which is considered a gold standard for concordance and accuracy.

### Ethical clearance

Ethical clearances from the respective institutions and national ethical clearance bodies have been secured for HDSS and CHAMPS activities. HDSS activities have standing approvals for continuing routine activities, including VA. All participants provided informed, voluntary, written consent. Consent was obtained from the responsible person in the family (the head of the household, the mother of the deceased child, or any eligible family member). Written informed consent was obtained from the parent/guardian of each participant under 18 years of ageTo keep anonymity and confidentiality, we did not share data that contained participants’ personal identifiers with any third party.

### Quality control

Individuals who completed at least a high school education and had experience working in the existing HDSS collected VA data. They received a two-week training on the questionnaires, recording, contacting close relatives, and data collection procedures. The training included sessions on discussing individual symptoms and their description in the local language for easy recognition by the respondents and demonstration of interviewing techniques by research team members. The field coordinators and supervisors continuously monitor data collection in the field to check progress and resolve problems that enumerators may have encountered during fieldwork.

### Data management and analysis

Data were analysed using STATA version 16. Means and standard deviations (SDs) were presented for continuous variables, medians and interquartile ranges for skewed variables, and counts and percentages for categorical variables. Demographic characteristics included age, gender, occupation, religion, and household size. Variables with more than 45% missing data were excluded.

We considered stillbirth as the absence of life after the viability of pregnancy (≥28 weeks of gestation) and before and during the baby’s delivery. Neonatal death was defined as a death in a live-born baby in the first 28 days of life. We further classified neonatal death into very early, early, and late neonatal death if the death occurred in the first 24 hours (day 0), 1-6 days, and 7 to 28 days, respectively [40]. Infant death was defined as a baby’s death after 28 days of life and before the first birthday, and child death as death before celebrating his/her 5^th^ year birthday [41].

Cause-specific mortality fractions (CSMF) for each surveillance site and CoD were computed by dividing the number of deaths due to specific causes assigned by either InterVA-4 and CHAMPS’s DeCoDe over the total number of deaths evaluated. The underlying causes of death from InterVA and DeCoDe were compared for agreement and pattern in assigning the diagnosis.

After the respective underlying causes of death that DeCoDe assigned were mapped and matched to its corresponding verbal autopsy standard, the agreement of both methods was evaluated using their concordance and accuracy of CSMF. We compared the CSMF of InterVA4 against DeCoDe using Lin’s Concordance Correlation Coefficient (LCC), [42] which was calculated using a user-defined command made for Stata – “Concord”[43].

The LCC determines how far the observed data deviate from the line of perfect concordance, a line at 45 degrees in a scatterplot. Lin’s coefficient increases in value as a function of the nearness of the data’s reduced major axis to the line of perfect concordance (the accuracy of the data) and of the tightness of the data about its reduced major axis (the precision of the data). The bias bias-correction factor shows how far the best-line of shift is from the perfect concordance. The program (“Concord”) produces the LCC by multiplying the “Pearson correlation coefficient, r” with the bias-correction factor. Whereas the “Pearson correlation coefficient, r” is the measure of precision, the bias-correction factor is for accuracy [43].

The LCC was stratified across surveillance sites, age classification, and enrolment location to evaluate the performance of InterVA4. The stratification of the group was according to the WHO 2016 VA instrument guideline [36]: children aged < 1 year and aged 1-4 years. Accuracy is the measurement of the validity of a measurement’s exact value or how close the predicted value obtained in data is to the true value. Precision is defined as the degree of reproducibility of using the same measurement or procedure to measure the degree of consistency of independent measurements of the same variable [44]. The interpretation of the LCC we used is < 0.8 is poor, 0.81-90 –as good, and > 0.9 is excellent [45]. We also used the same interpretation for accuracy and precision.

Furthermore, to complement LCC in measuring the agreement between InterVA4 and DeCoDe, a mortality fraction ratio was calculated by dividing the CSMF generated by the InterVA4 with the DeCoDe’s (InterVA4 CSMF/DeCoDe CSMF) by surveillance site and for specific CoDs at a 95% confidence interval generated using the Koopman method to identify whether the interpretation between the two methods was lower or higher than expected [46]. This statistical method produces “the Koopman asymptotic score interval” for the ratio of probabilities in two-by-two contingency tables and works well for small sample sizes. The purpose of calculating these CIs was not to demonstrate statistical significance but to identify whether the CSMF ratio between InterVA–4 and DeCoDe interpretations was significantly lower or higher than that expected from chance, considering the number of cases involved.

## Results

CHAMPS sites identified 7221 unique deaths (including stillbirths), of which 6,909 (95.7%) were enrolled from February 1, 2017, through December 30, 2021 (Figure 1). Of 6,909 enrolled deaths, 338 (4.9%) observations were removed from the analysis because they were missing CoDs generated from the InterVA-4 package of the Open-VA because of transcription errors, and 77 were removed because of a conflicting date of birth or death and CoDs. These deaths were also removed from the analysis. Of the remaining 6494 deaths, 2340 (36.0%) were stillbirths, 2321 (35.7%) were neonates, 967 (14.9%) were infants, and 866 (13.3%) were children aged 1-<5 years. Of these, 3641 deaths were excluded as they were not enrolled for MITS and only had InterVA-generated CoD. Therefore, we analyzed 2853 (43.9%) of 6494 deaths enrolled for MITS and subsequently had CoD information generated from both DeCoDe and VA.

**Figure 1.**
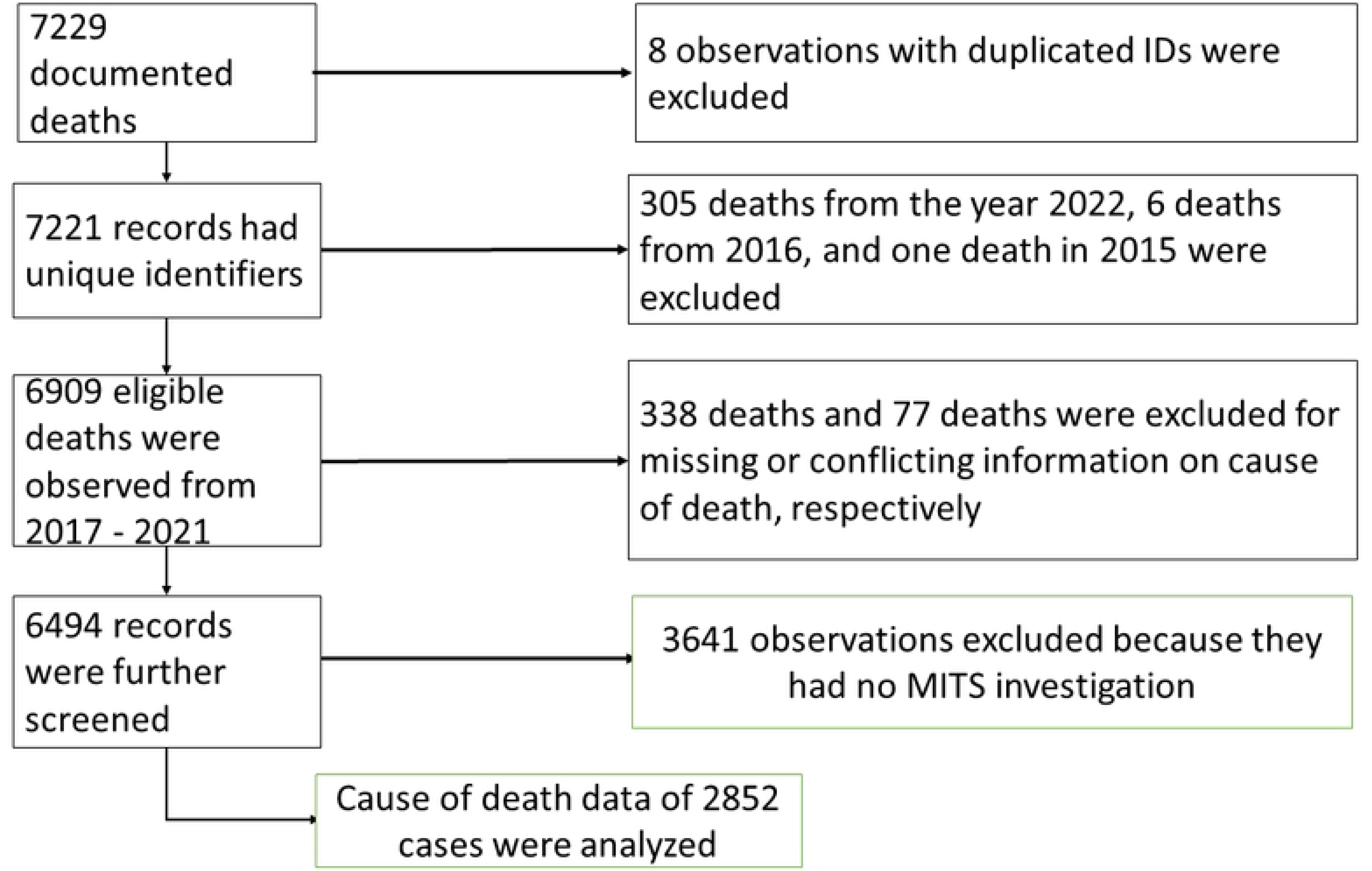

Of 2853 eligible deaths where both MITS and VA were undertaken, 1075 (37.7%) were stillbirths, 1077 (37.8%) neonatal deaths, 365 (12.8%) infant deaths, and 336 (11.9%) deaths of children aged 1-4 years. Around 30% (654) were enrolled from South Africa, 19% (545) from Kenya, 16.7% (476) from Mozambique, 12.2% (348) from Bangladesh, 11% from Sierra Leone (316) and Ethiopia (311), and 7% (203) from Mali. Across sites, the DeCoDe panel could not determine the CoDs for 78 (2.7%) of all deaths enrolled; of these, about half (40, 51.3%) were stillbirths, 15 (19.2%) were neonatal deaths, 15 (19.2%) infants, and 8 (10.3%) children aged 1-4 deaths (Table 1).

### Characteristics of study population and mortality groups across sites

Across sites, the mean age at death for U5 and newborns was 4 ± 10 months; females accounted for 44.1% of deaths (1251/2853), and more than half (55.1%) occurred in the dry season. The mean age of death was 4 (± 5.4) days for neonates, 5.4 (± 4) months for infants, and 2.2 (± 1.1) years. The mean gestational age for stillbirths was 34.3 weeks (95% CI 33.3, 35.6 weeks). Of 1075 recorded stillbirths, (207 (19.3%) were in Mozambique, 199 (18.5%) in Ethiopia, 177 (16.5%) in Bangladesh, 160 (14.9%) in Kenya, 153 (14.2%) in South Africa, 99 (9.2%) in Sierra Leone, and 80 (7.4%) in Mali. Overall, a large majority (89%) of deaths occurred in health facilities. About one-third of infant (111/365, 30.4%) and child deaths (102/336, 30.4%) occurred in the community. However, nearly all enrolled stillbirths (1037/1075, 96.5%) and neonatal deaths (1015/1077, 92.4%) occurred in health facilities (**Tables 1 and 2**).

The overall concordance of diagnoses across the surveillance sites and age groups was 0.75 (Table 2). The interVA4 method of assigning CoDs had better accuracy, but its precision compared to the DeCoDe was poor (<0.8). Stratified by surveillance sites, the overall concordance of all <5s deaths was lowest in Mali (0.64), and Ethiopia (0.83) and Mozambique sites (0.84) had good overall concordance.

Figure 2 shows the overall LCC of the CSMF generated by the InterVA4 against DeCoDe’s underlying causes of the death. We found the overall concordance level between the two systems to be poor, 0.75 (95%CI 0.73 – 0.76). The precision of the concordance was 0.98, while the accuracy was 0.76. The concordance coefficients were nearly the same across sexes for all CoDs and were higher for <5s enrolled at health facilities than those in the community. The determined CoDs for children aged < 1 year (0.69) were higher than those aged 1-4 years (0.28) despite their nearly no agreement when further stratified as stillbirths, neonates, and infants. However, the agreement considerably increased when those groups were combined (Figure 3).

**Figure 2.**
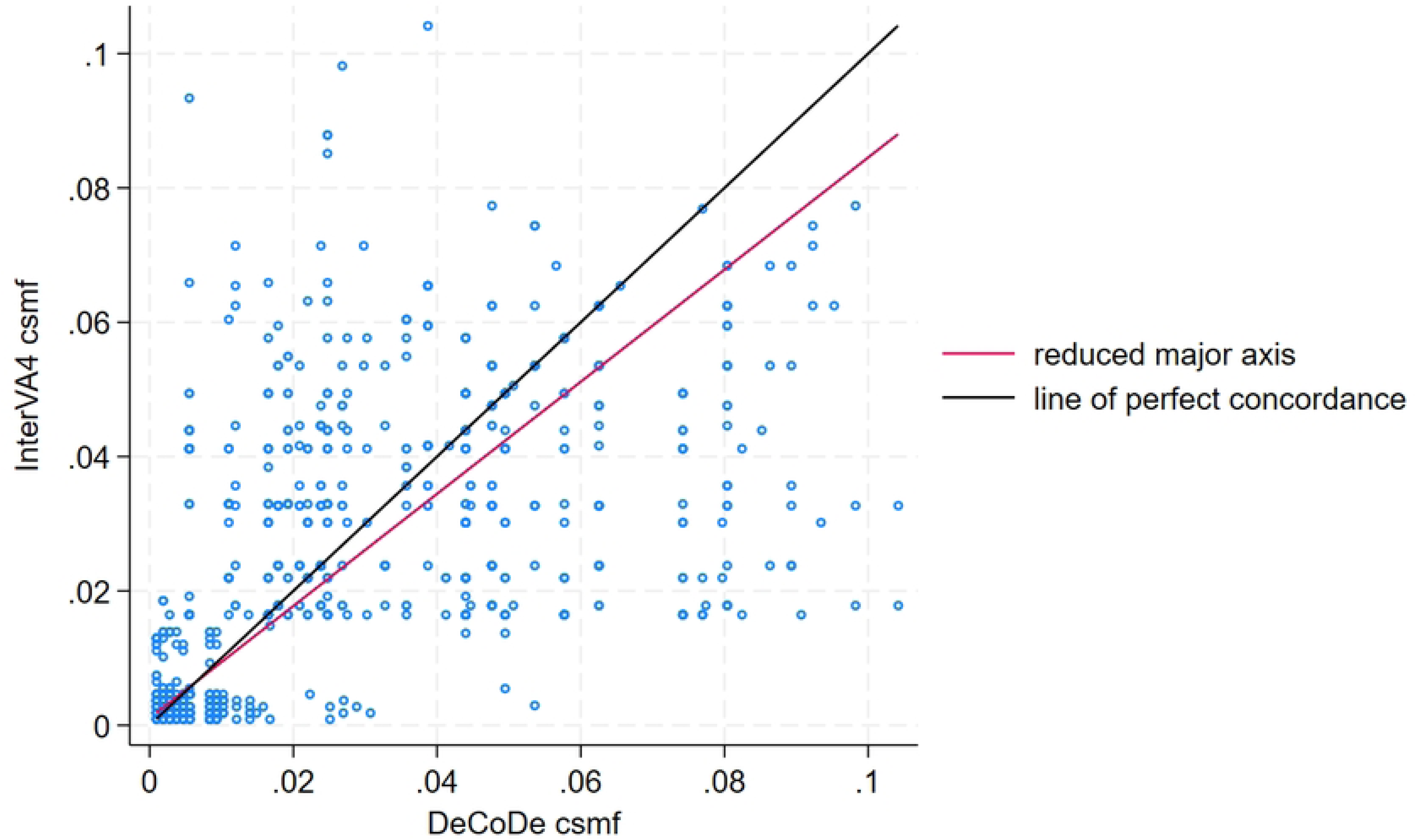

**Figure 3.**
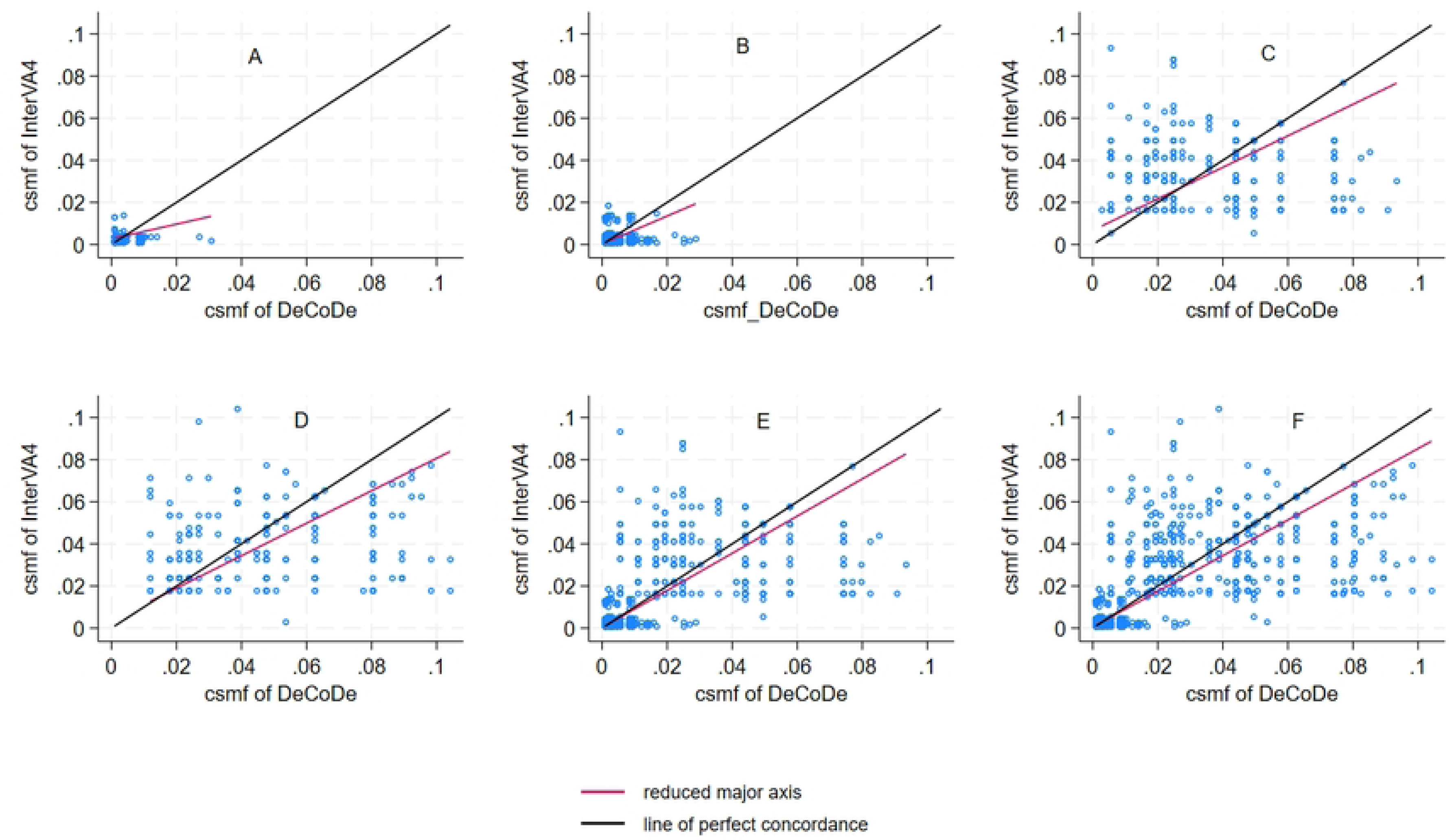

Cause-specific mortality fractions determined by InterVA4 and DeCoDe differed in important ways for some of the more common diseases (Table 3). In those surveillance sites where the DeCoDe panels determined HIV as the underlying CoD for some deaths, the InterVA4 model predicted considerably fewer HIV deaths, as demonstrated by CSMF. This pattern is also seen in many sites for diagnoses such as malnutrition (Ethiopia, Kenya, Mali, and Sierra Leone), neonatal sepsis (all sites except Mali), and birth asphyxia (all sites). However, the InterVA4 predicted a substantially higher proportion of deaths caused in most sites by prematurity (all sites, except Mali), malaria (Kenya, Mali, Mozambique, and Sierra Leone), malnutrition (Mozambique and South Africa), diarrheal diseases (except in Mozambique, which was lower), and meningitis (Kenya, Mozambique, Sierra Leone, and South Africa) than did DeCoDe.

Some CoDs were exclusively assigned by the InterVA4 model rather than the postmortem DeCoDe. For example, acute abdomen, renal failure, dengue fever, stroke, road and other traffic accidents, accidental falls, and exposure to the force of nature were exclusively assigned by the interVA4 model as the underlying CoDs and were not determined by the experts using postmortem MITS. Conversely, unspecified external causes of death, unspecified non-communicable diseases, congenital anomalies, digestive neoplasms, and unspecified neoplasms were exclusively determined as CoDs by experts using postmortem MITS. In addition, only the DeCoDe panels ascertained pulmonary tuberculosis as the CoD in South Africa. At the same time, the InterVA4 model did not predict it. The InterVA4 model exclusively assigned epilepsy in Ethiopia, Kenya, Mali, and Sierra Leone, but both InterVA4 and DeCoDe noted epilepsy as an underlying CoD in Mozambique and South Africa.

The InterVA4 CSMF and DeCoDe CSMF ratios were calculated with a 95% confidence interval using the CSMFs tabulated in Table 3 to show that many of these differences did not occur by chance. The InterVA4 CoDs of fresh and macerated stillbirths had the highest CSMF ratio, and unspecified neonatal CoD had the lowest CSMF ratio.

## Discussion

This study compared the InterVA4 model with experts’ determination of CoDs using advanced diagnostics and postmortem MITS and showed poor InterVA4 agreement and concordance in predicting the causes of death against DeCoDe among our <5s studied deaths. The concordance suffered from its accuracy (< 0.8), although the precision was good (>0.8).

Several other studies compared the InterVA4 with Physicians-Certified Verbal Autopsy (PCVA) and other standardized verbal autopsy diagnoses for public health equivalence to test its functionality and costs [16, 47–50]. Others have also studied the performance of InterVA4 with postmortem histologic findings. Knowing whether these tools lead to similar conclusions—and if not, how results differ--is important before relying on verbal autopsy-generated information as the general country-wide source of CoD and for planning and executing public health interventions [13]. This concept is particularly crucial in a setting without widespread mortality registrations and in resource-constrained areas.

Across surveillance sites, there were considerable differences in the two systems’ concordance, as Ethiopia and Mozambique’s LCC were good (>0.8) while the others were poor. Our findings could be explained by quality differences in collecting the VA data and the extent of CHAMPS’s concurrent utilization of VA data with other clinical information to assign the CoDs. The considerable agreement differences between children enrolled in health institutions and the community also substantiate the argument, as death enrolled in the health facility would have rich clinical information besides the VA compared to those cases enrolled in the community. Furthermore, other studies have reported that the extent and way of VA data collection determined how the InterVA4 assigned the respective CoDs [10, 16, 47, 51, 52]

However, we found the overall agreement in assigning the CoDs between the two systems to be poor. This finding is unsurprising as several studies also found significant differences between InterVA4 and PCVA or histologic findings [48, 53]. The concordance of InterVA4 considerably decreases for stillbirths, neonates, and infants at individual and population levels [33]. However, when they are combined, the level of agreement improves significantly. Most importantly, more than a quarter of the overall sample were stillbirths, where the InterVA4 is not designed to predict the causes of death. For example, most of the diagnoses assigned by the InterVA4 for stillbirths were VAs-11.01 or VAs-11.02. These assigned “macerated or fresh stillbirths” corresponded to the ICD-10 code of P95. In addition, InterVA4 did not assign congenital anomalies arising during the prenatal period, limiting its CoD equivalence compared to DeCoDe’s.

However, our findings did not agree with other studies that indicated an excellent concordance between the assigned causes of death between the InterVA4 model and several PCVA findings [47]. InterVA4 performed well in identifying malnutrition and certain perinatal conditions as the underlying CoDs, similarly to the DeCoDe. For example, the ratio of the proportion of malnutrition, birth asphyxia, and prematurity was closer to one or slightly higher, meaning better equivalence in assigning those conditions..

Furthermore, the DeCoDe captures the overall mortality chain from underlying, intermediate, and immediate causes of death, which is not done with InterVA4. In this study, we could only compare the InterVA4 models’ most likely underlying causes of death to the underlying causes attributed to DeCoDe. Comparing only the underlying CoDs may potentially limit the overall correlation of causes of death between the two approaches, as many deaths in live-born children occur after a complicated course of multiple causes [54]. For example, a neonate born prematurely could die of sepsis after admission to an intensive care unit; in this case, DeCoDe would account for both causes. The InterVA4, however, would mostly likely predict either of the causal chains, missing the overall causal chain. These complete causal-chain scenarios identified by the DeCoDe panel would be based on pieces of evidence from MITS and microbiological, clinical, and VA data. The DeCoDe process does involve clinical judgment in some cases, as attributing causes of death from multiple results can be complex, and clinical information, in particular, can be incomplete, incorrect, or absent [55]. Nonetheless, errors should be few as the procedure is designed to use the best possible set of information.

Another difference between the two methods is that the InterVA4 model mostly tends to assign the stillbirths – either fresh or macerated—underlying CoDs in 80 % of the cases, while the remaining CoDs designated were prematurity and intrauterine hypoxia; these CoDs, that the InterVA4 mainly assigned for stillbirths were VA-11.01 and VA-11.02 corresponds to the ICD code P95 – undetermined or unspecified causes of death. There were substantial differences in assigning ICD code P95 between the two methods unrelated to chance, as the InterVA4 assigned more than the DeCoDe. The differences could arise from the VA data quality, the algorithm design, or the MITS’s accuracy in determining the most likely causes of death.

Similarly, prematurity was also more often assigned by InterVA4 than the DeCoDe across sites, which could also be related to the level of certainty in determining the pregnancy’s gestational age, which the DeCoDe panel uses when assigning prenatal mortality. Moreover, those babies born prematurely are most likely to have impending birth asphyxia or respiratory distress, and the DeCoDe panel assigned more birth asphyxia than the InterVA4 model. These differences point out the relationship and the complexities of the causal chain that were responsible for the <5s deaths.

## Conclusion

Our findings point out that VA diagnosis alone, as generated by InterVA4, often incorrectly predicts causes of death among <5s, using DeCoDe findings as the gold standard. The InterVA4 model lacks precision in determining the underlying causes of death and cannot predict some conditions like congenital anomalies. Future improvement of the reliability and validity of VA data by strengthening the quality of data collection and automatically assigning CoDs using robust and new technologies, such as artificial intelligence, is recommended. Improving models to better predict causes of death, perhaps by using information from deaths that also have information from postmortem diagnostic assessments such as DeCoDe, would improve the usefulness of VA as a tool to inform health policies [56, 57].

Overall, the role of the VA as a tool for diagnosing and tracking the progress of mortality data among U5s is essential despite the noted shortcomings. Using the DeCoDe process that combines Minimally invasive tissue sampling (MITS) and other techniques could provide data to help improve CoDs determination. The data should subsequently be utilized to improve the CoD determination algorithms of VA and its diagnostic ability.

## Data Availability

The data is publicly available at https://doi.org/10.17605/OSF.IO/JSZNK

## Acknowledgments

We would like to thank all the community members from the CHAMPS sites and their local authorities for their support. We thank all the CHAMPS teams on the sites, including their legal representatives, for their support, the researchers, and the field staff involved in this project. We also thank the Program office of the CHAMPS project at Emory University for their continuous support and the local funders of each HDSS. Finally we would like to thank Zachary Madewell and Beth Tippett Barr for providing their feedback in data handling and drafting of the manuscript, respectively.

## Author contributions

NA, DB, and CY were involved in every stage of the study. All co-authors, drafted, commmented, edited, and approved submission.

## Ethics and consent

Ethical clearances from the respective institutions and national ethical clearance bodies have been secured for the HDSS and CHAMPS activities. The HDSS activities have standing approval for continuing activities, including VA. The ethical review committee of the respective Institutional Research Boards approved the study procedure at the sites. For all participants, informed, voluntary, written, and signed consent was obtained from the responsible person in the family (usually the head, the mother, or eligible family members). Written informed consent was obtained from the parent/guardian of each participant under 18 years of age. We did not share data containing participants’ identifiers with a third party to maintain confidentiality.

## Disclaimer

The findings and conclusions in this report are those of the author(s) and do not necessarily represent the views of the U.S. Centers for Disease Control and Prevention.

## Data availability statement

The data supporting this study’s findings are available on request from the corresponding author, [Nega Assefa].

## Supporting information

**S1 Figure:** Flowchart of deaths included in the analysis

**S2 Figure:** Concordance of cause-specific mortality fractions of the underlying causes of death between InterVA4 and DeCoDe for CHAMPS surveillance sites

**S3 Figure:** Concordance of cause-specific mortality fractions of the underlying causes of death between InterVA4 and DeCoDe for CHAMPS surveillance sites by age group.

